# Association of C-reactive protein and Interleukin-6 single nucleotide polymorphisms with preterm labour: results of a pilot study

**DOI:** 10.1101/2021.05.18.21257373

**Authors:** Akira Sato, Hiroshi Miura, Yohei Onodera, Hiromitu Shirasawa, Saeko Kameyama, Akiko Fujishima, Yuko Kikuti, Mayumi Goto, Toshiharu Sato, Yukihiro Terada

## Abstract

Preterm labour (PTL) can be described as an inflammatory event. C-reactive protein (CRP) and Interleukin-6 (IL-6) are key members of the innate immune response that play major roles in inflammation. The main objective of this case-control study was to determine the association of specific CRP and IL-6 polymorphisms with PTL. The study was carried out in a total of 31 Japanese women with PTL and 28 control women with normal pregnancy. Four SNPs in the CRP gene (rs1800947, rs3091244, rs2794521, and rs3093059) and two SNPs in the IL-6 gene (rs2097677 and rs1800795) were genotyped using a polymerase chain reaction-restriction fragment length polymorphism (PCR-RFLP) assay. Biochemical parameters were assayed and cervical length measurements were performed with ultrasound sonography. There were no significant differences in inflammatory markers, including white blood cell count, CRP, neutrophil elastase in cervical mucous, and foetal fibronectin in vaginal discharge, between the PTL and control groups. The frequency of rs1800947 minor allele (G) was significantly higher in the PTL group than in the control group. This finding has not been previously reported. We suggest that mutations in rs1800947 may lead to PTL. Thus, the rs1800947 SNP may be useful as a genetic marker for PTL risk assessment in pregnant women.

## Introduction

Preterm birth remains the major cause of neonatal morbidity and mortality. PTL may have several causes. One of the most convincing hypotheses is that, as in full-term labour[1], PTL may be the result of an inflammatory reaction. Before the onset of labour, a pro-inflammatory condition can be induced in reproductive tissues such as the myometrium, cervix, and decidua. This condition is characterized by pro-inflammatory cytokines such as IL-6 and -8, and the infiltration of monocytes and macrophages. Presumably, the inflammatory reaction may play a role in the transition of the myometrium from a quiescent to a contractile state. Infection as well as uterine hyperextension and bleeding in the contractile state could trigger the pathological contraction of the uterus, leading to PTL.

C-reactive protein (CRP), which plays an important role in innate immunity, is an acute reaction-related protein produced by the liver in response to IL-6. A genome-wide association study in Japanese subjects revealed the association between serum CRP levels and a single nucleotide polymorphism (SNP) in the promoter region of IL-6 [2]. Meanwhile, CRP polymorphisms have been associated with various kinds of inflammatory diseases, such as bacterial infections [3,4], heart disorders [5–10], and malignant tumours [11–14]. Specific SNPs have previously been linked to preterm birth and labour [15,16]. However, there are only a few reports on the association between IL-6 SNPs and PTL [17–19], and to the best of our knowledge, none on the association between CRP SNPs and PTL. Our attentions were payed to four SNPs in the CRP gene (rs1800947, rs3091244, rs2794521, and rs3093059) and two SNPs in the IL-6 gene (rs2097677 and rs1800795) in the reason why rs1800947, rs3091244, and rs2794521 associated with CRP level in bacterial infection [4], rs2097677 and rs3093059 associated with serum CRP level in a genome-wide association study in Japanese [2], and IL-6 rs1800795 associated with preterm birth [19] were reported previously.

Thus, the purpose of this study was to examine the association of specific CRP and IL-6 polymorphisms with PTL. Our findings suggest that mutations in rs1800947, a CRP SNP, may lead to PTL. Based on these findings, we propose that rs1800947 could be used as a genetic marker for PTL risk assessment in pregnant women.

## Materials and Methods

### Ethics statement and study subjects

The study protocol was approved by the institutional ethics committee of Akita University of Medicine (Approval number: 1431) and written informed consent was obtained from all participants.

This study was conducted in Akita prefecture at Akita University Hospital between March 2017 and December 2019. This is a case-control study carried out in a total of 31 Japanese women with PTL and 28 control women with normal pregnancy. The diagnosis of PTL was based on clinical criteria of regular uterine contractions accompanied by a change in cervical dilation, effacement, or both, or initial presentation with regular contractions and cervical dilation of at least 2 cm at 22–36 weeks of gestation [20]. Blood sampling, cervical and vaginal sampling, and transvaginal ultrasound examination were performed when PTL was diagnosed. In the control group, these samplings and measurements were performed at 26–28 weeks of gestation on periodical medical examination.

### Genotyping analysis

Fresh blood was collected in EDTA tubes and subjected to density gradient centrifugation using Leuco Spin Medium (pluriSelect Life Science, Leipzig, Germany). After samples were centrifuged for 30 minutes at 1000g at room temperature, the layer corresponding to peripheral blood mononuclear cells (PBMCs) was collected and stored at -80 °C until use for SNP analysis. Genomic DNA was extracted from PBMCs using QIAamp DNA Mini Kit (QIAGEN, Germany). Four SNPs in the CRP gene (rs1800947, rs3091244, rs2794521, and rs3093059) and two SNPs in the IL-6 gene (rs2097677 and rs1800795) were genotyped in all samples using a polymerase chain reaction-restriction fragment length polymorphism (PCR-RFLP) assay. The primer sequences are shown in Table 1. PCR reactions were performed in a total volume of 50 µl containing 50 ng genomic DNA, 3 µl of each primer (0.6 µM), 200 µM of each dNTP, 10× PCR buffer for KOD-Plus-Ver.2 (TOYOBO, Japan), 1.5 mM MgSO4 and 1.0 U of KOD-Plus- (TOYOBO, Japan). The amplification was carried out with an initial denaturation step at 94 °C for 2 min, followed by 30 cycles of 30 seconds at 55 °C and 60 seconds at 68 °C. An ABI Prism 3130xl Genetic Analyzer (Applied Biosystems) was used to sequence the PCR products. BEX Co., Ltd. cooperated in the genotyping of these SNPs throughout the whole process.

**Table1.**
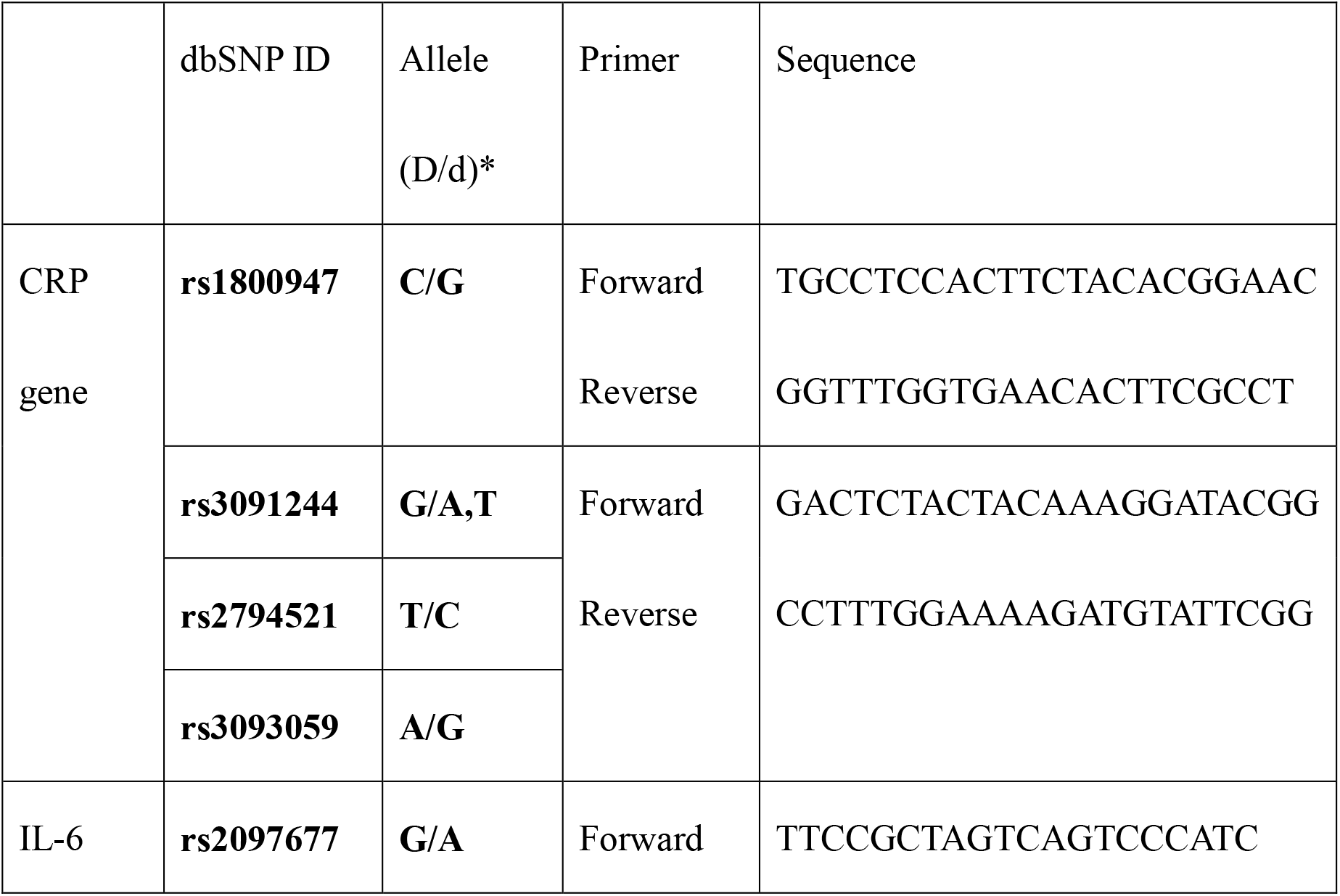
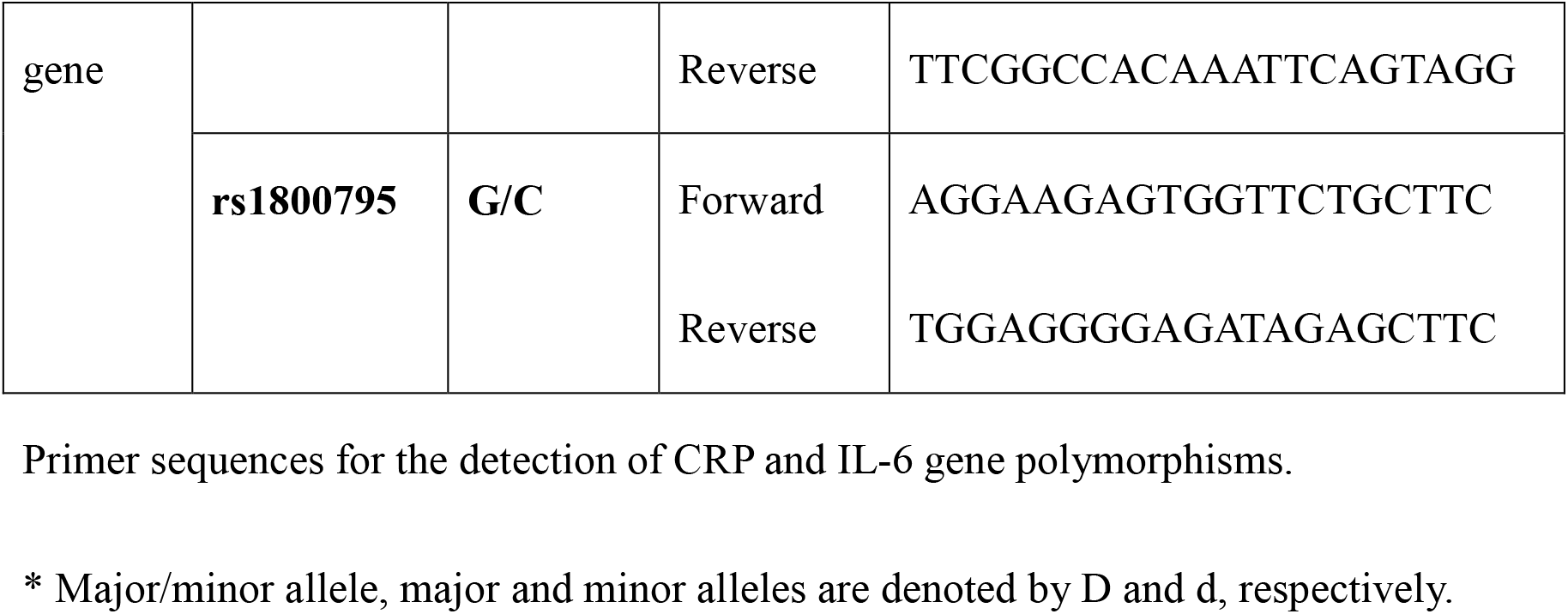
Primer sequences for the detection of CRP and IL-6 gene polymorphisms.

### Biochemical parameters and cervical length measurements

Serum CRP levels and white blood cell counts (WBC) were obtained in the main laboratory by a standard laboratory method. The samples of cervical fluid were obtained with cotton swab placed in the endocervical canal and were mixed and diluted with phosphate buffer, and the extracted buffer was filtrated. The concentration of neutrophil elastase (NE) in sample fluid was determined using a latex agglutination immunoassay kit (Sanwa Kagaku Co., Nagoya, Japan)[21]. Samples of foetal fibronectin (FF) were obtained by taking a cotton swab from posterior vaginal fornix. The concentration of FF in the sample fluid was determined using an enzyme-linked immunosorbent assay kit (Sekisui Medical Co., Tokyo, Japan)[22]. The cut-off points of NE and FF were 1.60 µg/mL and 50 ng/ml, respectively. SRL Co., Ltd. cooperated on NE and FF measurements. All cervical length measurements were carried out according to the guidelines of the Foetal Medicine Foundation [23]. Immediately after delivery, 1–2 ml blood from an umbilical cord artery was taken in a heparin syringe and examined with the laboratory blood gas analyser (ABL80FLEX, RADIOMETER Co., Japan).

### Statistical analysis

All data were analysed using the statistical software package JMP 12.2.0 (SAS Institute Inc, Cary, NC, USA). Continuous variables are presented as means ± standard deviations or median (interquartile range) and categorical variables are presented as fraction. Univariate analysis used student’s t-test, Wilcoxon-rank sum test (if variable was not normally distributed), Chi squared test, or Fisher’s exact test as appropriate. Normality was assessed using the Shapiro-Wilk test. Odds ratios with 95% confidence intervals for the genotypes were calculated using Chi squared test, or Fisher’s exact test. All tests are two sieded and p values less than 0.05 were considered statistically significant. Genotype frequencies of all SNPs were tested for Hardy-Weinberg equilibrium (HWE) by chi square test.

## Results

Demographic and clinical parameters of the PTL and control groups are shown in Table 2. A significant difference in cervical length was observed between the PTL and control groups (p<0.001). In contrast, no significant differences in CRP levels were observed between the two groups. The frequencies of positive NE and FF were not significantly different between the two groups.

**Table 2.** Comparison of demographic and clinical parameters between the PTL and Control groups.

|  | PTL<br>(N=31) | Control<br>(N=28) | p-values |
| --- | --- | --- | --- |
| Maternal age (years) | 33.0±4.92 | 33.9±5.28 | 0.486 |
| Primiparous/Multiparous | 15/16 | 11/17 | 0.601 |
| Primigravid/Multigravid | 22/9 | 15/13 | 0.189 |
| History of premature<br>delivery | 5/26 | 4/24 | 1.000 |
| GA at examination<br>*(weeks) | 27(26-28) | 27(26-27) | 0.180 |
| WBC (/μL) | 7958±1447 | 8689±1914 | 0.101 |
| CRP (mg/dL) | 0.15(0.08-0.33) | 0.13(0.06-0.22) | 0.347 |
| Cervical length (mm) | 27.5±9.81 | 49.2±11.4 | < 0.001 |
| PMNE<br>(negative/positive) | 15/16 | 14/14 | 1.000 |
| Fibronectin | 29/2 | 28/0 | 0.492 |
| (negative/positive) |  |  |  |

\*GA at examination: gestational age at blood and vaginal sampling and ultrasound measurement. Maternal age, WBC, and Cervical length are presented as means± standard deviation. GA at examination and CRP are presented as median (interquartile range). All categorical variables are presented as fraction.

Perinatal and neonatal parameters are presented in Table 3. Gestational age at delivery was significantly lower in the PTL group (p<0.05). Mode of delivery, neonatal body weight, sex, Apgar Score, and pH of the umbilical artery were not significantly different between the two groups.

**Table 3.** Comparison of perinatal and neonatal data

|  | PTL<br>(N=31) | Control<br>(N=28) | p-values |
| --- | --- | --- | --- |
| GA at delivery<br>(weeks) | 37(36-37) | 39(38-40) | < 0.001 |
| Mode of delivery<br>(vaginal/cesarean) | 21/10 | 18/10 | 0.790 |
| Neonatal Weight | 2792(2542-3022) | 2895(2752-3086) | 0.054 |
| Male/Female | 20/11 | 11/17 | 0.069 |
| Apgar Score |  |  |  |
| 1 minutes |  |  |  |
| 0–3 | 2 | 0 | 0.742 |
| 4–6 | 1 | 1 |  |
| 7–10 | 28 | 27 |  |
| 5 minutes |  |  |  |
| 0–3 | 0 | 0 |  |
| 4–6 | 1 | 0 | 1.000 |
| 7–10 | 30 | 28 |  |
| pH of Umbilical Artery | 7.31±0.067 | 7.32±0.057 | 0.676 |

\*GA at examination: gestational age at delivery.
GA at delivery and pH of Umbilical artery are presented as means± standard deviation.
Neonatal weight is presented as median (interquartile range). All categorical variables are presented as fraction.

Genotype and allele distribution of candidate SNPs are described in Table 4. Rs1800947 in the PTL group (MAF0.19, p<0.001) and rs3091244 in both the PTL (MAF0.22, p<0.001) and control (MAF0.28, p<0.001) groups were deviated from HWE, while the other SNPs were in HWE in both groups. All participants of both groups had GG genotype of IL-6 rs1800795. A significant difference was observed in the genotype distribution of rs1800947 between the PTL and control groups. Specifically, the frequency of the minor G allele was significantly higher in the PTL group (p<0.05).

**Table 4.** Genotypes and alleles distribution of CRP rs1800947, rs3091244, rs2794521, and 3093059, and IL-6 rs2097677, and rs1800795 polymorphisms in the PTL and Control groups.

| SNPs (D/d) * | PTL(N=31) | Control(N=28) | OR(95%CI) | p-values |
| --- | --- | --- | --- | --- |
| rs1800947 | HWE | HWE |  |  |
| (C/G) | p<0.001 | p=1.000 |  |  |
|  | MAF 0.19 | MAF 0 |  |  |
| CC | 27 | 28 | 1.000(Ref) |  |
| CG | 2 | 0 | - | 0.157 |
| GG | 2 | 0 | - | 0.157 |
| CG+GG | 4 | 0 | - | 0.049 |
| Allele |  |  |  |  |
| C | 56 | 56 | 1.000(Ref) |  |
| G | 6 | 0 | - | 0.017 |
| rs3091244 | HWE | HWE |  |  |
| (G/T, A) | p<0.001 | p<0.001 |  |  |
|  | MAF 0.22 | MAF 0.28 |  |  |
| GG | 17 | 15 | 1.000(Ref) |  |
| GT | 9 | 8 | 0.993(0.305-3.226) | 0.990 |
| GA | 5 | 2 | 0.453(0.076-2.690) | 0.376 |
| TT | 0 | 2 | - | 0.145 |
| AT | 0 | 1 | - | 0.295 |
| GT+GA+TT+AT | 14 | 13 | 0.950(0.341-2.650) | 0.922 |
| Allele |  |  |  |  |
| G | 48 | 40 | 1.000(Ref) |  |
| T | 9 | 13 | 0.577(0.227-1.674) | 0.252 |
| A | 5 | 3 | 0.720(0.162-3.200) | 0.665 |
| rs2794521<br>(T/C) | HWE<br>p=0.230<br>MAF 0.17 | HWE<br>p=0.604<br>MAF 0.08 |  |  |
| TT | 20 | 23 | 1.000(Ref) |  |
| TC | 11 | 5 | 0.395(0.117-1.332) | 0.128 |
| Allele |  |  |  |  |
| T | 51 | 51 | 1.000(Ref) |  |
| C | 11 | 5 | 0.455(0.147-1.402) | 0.163 |
| rs3093059<br>(A/G) | HWE<br>p=0.344<br>MAF 0.14 | HWE<br>p=0.603<br>MAF 0.23 |  |  |
| AA | 22 | 17 | 1.000(Ref) |  |
| AG | 9 | 9 | 0.773(0.251-2.368) | 0.652 |
| GG | 0 | 2 | - | 0.119 |
| AG+GG | 9 | 11 | 0.632(0.214-1.871) | 0.406 |
| Allele |  |  |  |  |
| A | 53 | 43 | 1.000(Ref) |  |
| G | 9 | 13 | 0.562(0.219-1.438) | 0.226 |
| rs2097677<br>(G/A) | HWE<br>p=0.076<br>MAF 0.24 | HWE<br>p=0.890<br>MAF 0.17 |  |  |
| GG | 16 | 19 | 1.000(Ref) |  |
| GA | 15 | 8 | 0.449(0.152-1.330) | 0.145 |
| AA | 0 | 1 | - | 0.364 |
| Allele |  |  |  |  |
| G | 47 | 46 | 1.000(Ref) |  |
| A | 15 | 10 | 0.681(0.278-1.671) | 0.400 |

\*Major/minor allele, major and minor alleles are denoted by D and d, respectively.
OR: Odds ratio. CI : Confidence interval. HWE: Hardy-Weinberg equilibrium. MAF:
Minor allele frequency.
All participants of both groups had GG genotype of IL-6 rs1800795.

## Discussion

In this study, we did not find significant differences in WBC and serum CRP levels, which are common markers of inflammation and bacterial infection, between the PTL and control groups. These results are consistent with the fact that pregnant women experiencing PTL often show no signs of clinical inflammation or infection. Moreover, no significant differences in NE and FF, which are local inflammatory markers in the uterus, cervix, and vagina, were found between the two groups. Previous reports have already indicated that their efficacy as PTL markers is controversial [24-27]. Regarding perinatal prognosis, the gestational age at delivery was significantly lower in the PTL group than in the control group.

The examination of four candidate SNPs in the CRP gene and two candidate SNPs in the IL-6 gene that are associated with CRP production indicated that the frequency of rs1800947 minor G allele in the CRP gene was significantly higher in the PTL group. The human CRP gene is located on chromosome 1q21–23, spanning approximately 1.9 kb and containing two exons. Rs1800947 is a synonymous coding polymorphism located on exon 2 of the CRP gene, which was reported for the first time by Cao et al. [28]. Subsequently, various diseases have been associated with rs1800947, such as coronary artery disease [5,6], myocardial infarction [7–10], cerebrovascular ischemia-related disease [29–31], prostate cancer [12,13], colorectal cancer [14], diabetes [32], and depression [33]. The association of this SNP with low CRP levels was also described in myocardial infarction [7], coronary artery bypass surgery [6], coronary artery disease [5], cerebrovascular ischemia-related disease [31], diabetes [32], prostate cancer [12], and oesophageal cancer [34]. In this study, we could not statistically examine CRP levels according to the genotype of each SNP because of the limited number of study subjects.

CRP was discovered as a protein bound to phosphocholine, which is a kind of phospholipid present in the pneumococcal surface. CRP is a representative acute reaction-related protein that increases in the blood during inflammation, infection, and tissue injury. When CRP binds to various ligands such as phosphocholine or polycationic molecules, the classical complement pathway can be activated. Furthermore, CRP reinforces the production of inflammatory cytokines and tissue factors (TFs), and the expression of plasminogen activator-1 (PAI-1) in macrophages, smooth-muscle cells, and vascular endothelial cells.

According to previous reports, mutations in CRP rs1800947 could be involved in the occurrence and development of diseases because CRP plays a key role in innate immunity. Multiple factors such as environmental agents could induce these diseases in the presence of a particular SNP mutation. Given our results, we hypothesize that the SNP mutation in the CRP gene could reinforce the production of chemical mediators such as IL-6 by macrophages, which may lead to a pro-inflammatory state in the myometrium and decidua that induces labour.

IL-6 is produced by monocytes, macrophages, and endothelial cells. The relationship between IL-6 and PTL has been extensively described [35-47]. IL-6 increases the expression of myometrial oxytocin receptor and promotes oxytocin secretion, increasing the reactivity of oxytocin [1, 48]. Thus, IL-6 may initiate the pathogenesis of PTL. In this study, we did not observe significant differences in the frequencies of alleles in the IL-6 rs2097677 and rs1800795 SNPs. Adam et al. reported no significant impact of rs1800795 on preterm labour with rupture [17]. Felix et al. reported no significant association of rs1800795 with preterm birth in a study of obstetrical complications in 1,626 pregnant women [18]. Conversely, Digna et al. described rs1800795 as one of the significantly relevant gene sites in association with preterm birth [19]. In this study, the results on rs2097677 and rs1800795 were consistent with the former two reports.

One limitation of this study was the deviation of rs1800947 from HWE, which indicated the possibility of structured subjects in this study. Other limitations include a small sample size, the lack of serum IL-6 measurements, and the absence of pathological examination of the placenta. A prospective observational study should be conducted and will be necessary to increase the number of subjects in the future.

## Conclusion

An association between the rs1800947 minor G allele and PTL was observed in this study. Given the association of rs1800947 with other diseases, we suggest that PTL may occur as a result of rs1800947 gene mutation. Thus, rs1800947 could be used as a genetic marker for PTL risk assessment in pregnant women.

## Data Availability

All data are fully available without restriction.

## Acknowledgments

The authors thank the participants and staff for their participation in the study. We would like to thank Editage (www.editage.com) for English language editing.

